# Methylome-wide studies of six metabolic traits

**DOI:** 10.1101/2024.05.29.24308103

**Authors:** Hannah M. Smith, Hong Kiat Ng, Joanna E. Moodie, Danni A. Gadd, Daniel L. McCartney, Elena Bernabeu, Archie Campbell, Paul Redmond, Adele Taylor, Danielle Page, Janie Corley, Sarah E. Harris, Darwin Tay, Ian J. Deary, Kathryn L. Evans, Matthew R. Robinson, John C. Chambers, Marie Loh, Simon R. Cox, Riccardo E. Marioni, Robert F. Hillary

## Abstract

Exploring the molecular correlates of metabolic health measures may identify the shared and unique biological processes and pathways that they track. Here, we performed epigenome-wide association studies (EWASs) of six metabolic traits: body mass index (BMI), body fat percentage, waist-hip ratio (WHR), and blood-based measures of glucose, high-density lipoprotein (HDL) cholesterol, and total cholesterol. We considered blood-based DNA methylation (DNAm) from >750,000 CpG sites in over 17,000 volunteers from the Generation Scotland (GS) cohort. Linear regression analyses identified between 304 and 11,815 significant CpGs per trait at P<3.6×10^-8^, with 37 significant CpG sites across all six traits. Further, we performed a Bayesian EWAS that jointly models all CpGs simultaneously and conditionally on each other, as opposed to the marginal linear regression analyses. This identified between 3 and 27 CpGs with a posterior inclusion probability ≥ 0.95 across the six traits. Next, we used elastic net penalised regression to train epigenetic scores (EpiScores) of each trait in GS, which were then tested in the Lothian Birth Cohort 1936 (LBC1936; European ancestry) and Health for Life in Singapore (HELIOS; Indian-, Malay- and Chinese-ancestries). A maximum of 27.1% of the variance in BMI was explained by the BMI EpiScore in the subset of Malay-ancestry Singaporeans. Four metabolic EpiScores were associated with general cognitive function in LBC1936 in models adjusted for vascular risk factors (Standardised β_range_: 0.08 – 0.12, P_FDR_ < 0.05). EpiScores of metabolic health are applicable across ancestries and can reflect differences in brain health.

## Introduction

Measures of adiposity and lipids are central to profiling metabolic health. There are several clinical measures of metabolic health, which include body mass index (BMI), body fat percentage, waist-hip ratio (WHR), blood glucose levels, high-density lipoprotein (HDL) cholesterol, and total cholesterol. These traits have routinely been linked to health-related risks including cardiovascular disease (1–3), myocardial infarction (4), and stroke (2, 3, 5). Multiple associations between metabolic traits and cognitive function and rate of cognitive decline have also been observed (6–12). BMI is a widely assessed indicator of metabolic health but is limited by its inability to directly track the amount or distribution of fat in the body (13, 14). BMI has previously shown low specificity in identifying individuals with excess body fat (15). Considering multiple measures that track different aspects of adiposity (and related traits) may provide a more complete assessment of metabolic health. Furthermore, exploring the molecular correlates of these metabolic indices may help to inform the shared and unique biological processes and pathways that they are associated with.

The epigenetic modification DNA methylation (DNAm) is dynamic, tissue/cell-type specific, and can be affected by genetic and environmental factors. Epigenome-wide association studies (EWASs) have detailed associations between individual blood-based DNAm loci (CpG sites) and metabolic traits including BMI, WHR, HDL cholesterol, and total cholesterol (16–32). In our previous work, penalised regression models have been applied to DNAm data to develop molecular predictors for a multitude of complex traits. These epigenetic scores, or EpiScores, may augment associations with health outcomes when combined with their measured phenotypic counterparts (33–35). For example, an EpiScore for BMI increased the amount of variance in metabolic health outcomes accounted for by measured BMI alone by an average of 3% (36). An EpiScore for WHR was also associated with all-cause mortality in the same population of healthy older adults after adjusting for measured WHR (33).

Here, we modelled EWASs with both linear regression and Bayesian penalised regression on six metabolic traits in the Generation Scotland (GS) study (*N* > 17,000). In the former approach, we obtained marginal estimates for each CpG, which do not take into account correlations across CpGs. By contrast, the Bayesian penalised regression estimated CpG effects jointly so that the effect of each CpG was conditional on all other loci. We compared findings from the individual EWASs to determine whether the six traits showed unique or common methylomic signatures. We then trained EpiScores for the six metabolic traits in GS (*N* > 17,000) and projected them into two independent test cohorts – the Lothian Birth Cohort 1936 (LBC1936) and the Health for Life in Singapore (HELIOS) cohort. Finally, we tested metabolic trait EpiScore associations with general cognitive function level and change in LBC1936 (N *=* 861). Associations identified between EpiScores for metabolic traits and cognitive phenotypes could offer new opportunities to examine the relevance of metabolic health indicators to ageing, and cognitive and neurological health outcomes. A visual summary of the study is shown in **Figure 1**.

**Figure 1:**
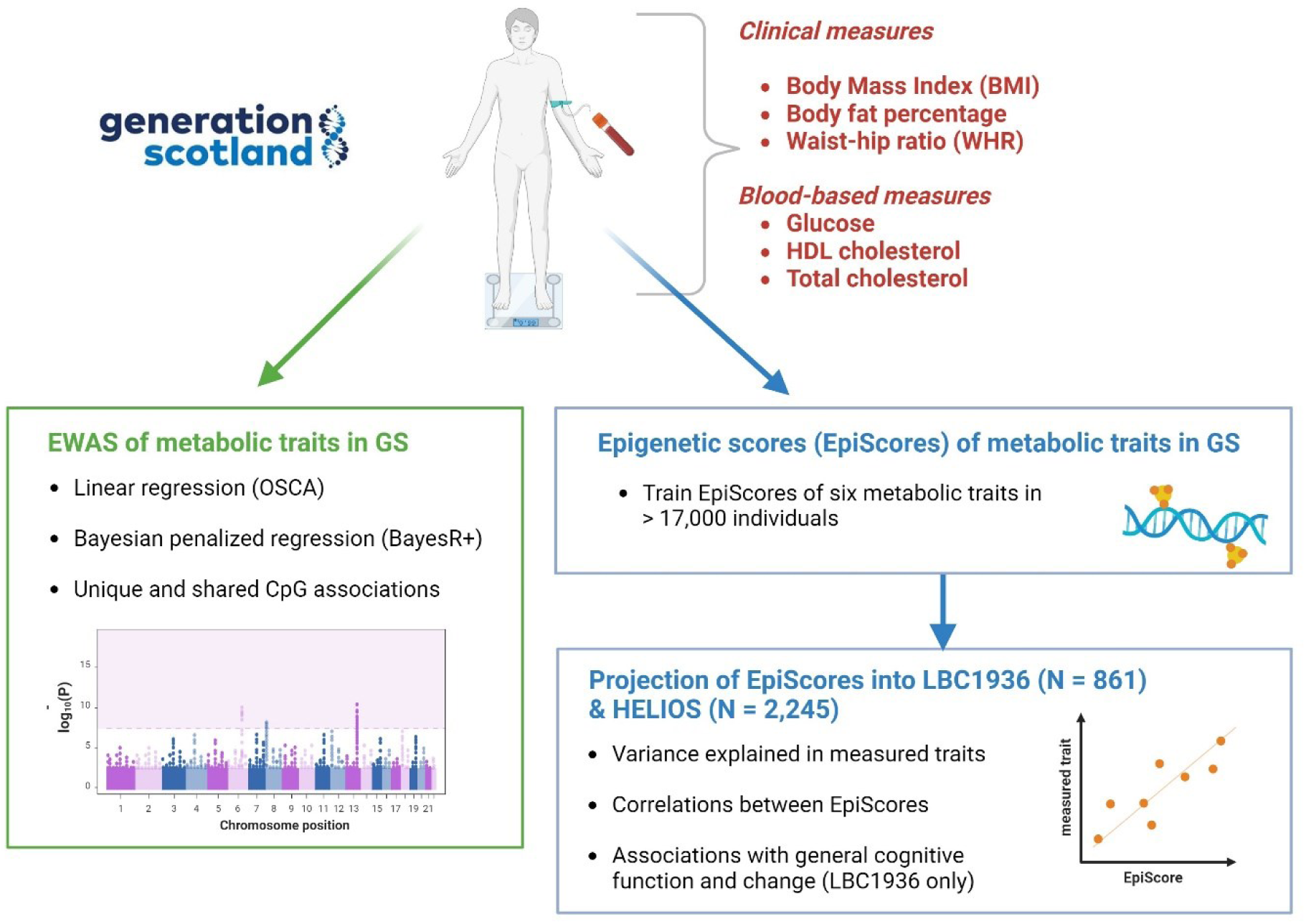
Summary of metabolic trait study. This figure provides an overview of the analysis performed in this study. Created with BioRender.com.

## Methods

### Generation Scotland Cohort

The Generation Scotland (GS) Cohort has been described in detail previously (37). Briefly, it is a Scotland-wide, family-based study of health. In the current study, 18,411 individuals had DNA methylation profiled on the Illumina EPIC array from blood samples taken at the study baseline between 2006 and 2011. Quality control (QC) details can be found in **Supplementary Methods**. 59% of the cohort was female and the mean age at baseline was 47.5 years (SD: 14.9). Six metabolic measures from GS were utilised in this study: body mass index (BMI, kg/m^2^), body fat percentage, waist-hip ratio (WHR), glucose (mmol/L), serum HDL cholesterol (mmol/L), and serum total cholesterol (mmol/L) (**Table 1**, **Supplementary Methods**).

**Table 1:**
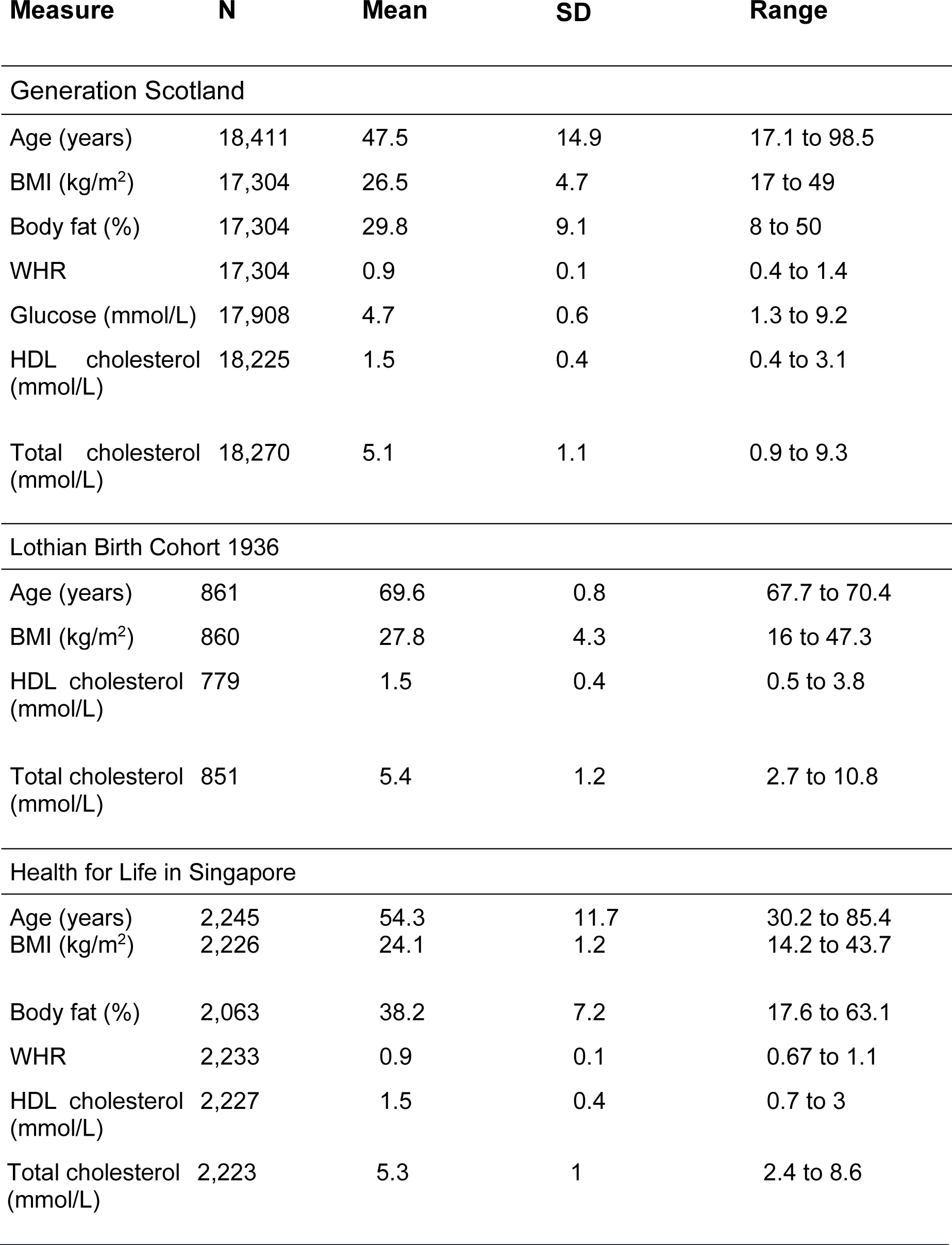
Cohort demographics for Generation Scotland, the Lothian Birth Cohort 1936 and the Health for Life in Singapore study. Table 1 shows the demographics of the data included in this study including N, mean, range and standard deviation for each variable after outlier removal. SD = standard deviation, BMI = body mass index, WHR = waist-hip ratio, HDL = high-density lipoprotein.

### The Lothian Birth Cohort 1936

The Lothian Birth Cohort 1936 (LBC1936) is a longitudinal study of ageing (38, 39). The study consists of individuals born in 1936, most of whom sat a general cognitive ability test at a mean age of 11 years in Scotland. Individuals living in the Lothian area were recruited to the LBC1936 study at around age 70 (baseline N=1,091). The volunteers undertook triennial testing across five waves of follow-up (ages ∼70, 73, 76, 79, and 82). Of those with blood-based DNA methylation data (profiled on the Illumina 450k array) at wave 1, the mean age was 69.6 years (SD: 0.83) with 49.4% females. QC and pre-processing for the DNA methylation in the LBC1936 can be found in **Supplementary Methods**. Three metabolic measures were utilised in this study: BMI (kg/m^2^), serum HDL cholesterol (mmol/L), and serum total cholesterol (mmol/L) (**Table 1**, **Supplementary Methods**). Thirteen cognitive tests were assessed longitudinally (details in **Supplementary Methods**).

### The Health for Life in Singapore cohort

The Health for Life in Singapore (HELIOS) study is a single-centre, multi-ancestry cohort of approximately 10,000 individuals residing in Singapore. A subset of the cohort in which Illumina EPIC DNA methylation data have been profiled has a mean age of 54.3 (SD: 11.7) and 61.2% of the cohort was female. The subset is made up of three self-reported ancestry groups: Chinese and other East Asian (Chinese) (N = 1,778), Malay and other South-East-Asian (Malay) (N = 242), and South Asian (Indian and other countries from the Indian subcontinent) (N = 225). QC and pre-processing of DNA methylation in HELIOS can be found in the **Supplementary Methods**. Five metabolic measures were utilised in this study: BMI (kg/m^2^), body fat percentage, WHR, serum HDL cholesterol (mmol/L) and serum total cholesterol (mmol/L) (**Table 1**, **Supplementary Methods**).

### Epigenome-wide association studies of six metabolic traits in GS

Linear regression models tested for associations between 752,722 CpG sites and each of the six metabolic traits in GS using the fast linear method in the OmicS-data-based Complex trait Analysis (OSCA) software (40). To facilitate less computationally expensive analyses, phenotypes were regressed on age, age^2^, sex and family structure (to account for relatedness in GS) (41)) using linear mixed-effects models (lmekin function from the coxme package (version: 2.2.18.1) in R) (42). Family structure was modelled with a kinship matrix constructed using the R package kinship2 (version: 1.9.6). CpG M-values were pre-corrected for age, sex and experimental batch (*N =* 121 batches) in linear regression models using the lm function in R. Residuals from the regression models for each outcome trait and CpG were taken forward for the EWASs. An epigenetic smoking score, EpiSmokEr, derived using the SSc method which adds up methylation levels of 187 CpG sites found to be significantly associated with smoking in a study by Zeilinger *et al* (43, 44) and Houseman-estimated white blood cell proportions (45) were included as fixed-effect covariates in the OSCA analysis. Finally, the first 20 methylation principal components (PCs) were included as covariates to account for potentially unmeasured confounders. Descriptive statistics can be found in **Supplementary Table 1**. A significance level of P < 3.6 × 10^-8^ was set to detect significantly associated CpGs as suggested by Saffari *et al* in a study investigating significance thresholds in EWAS using a simulation approach (46). Mapping of CpG sites to genes was performed using Illumina annotation files. Principal component analyses (PCA) were performed on the significantly associated CpG sites from each metabolic trait EWAS. The number of approximate independent signals was denoted as the cumulative number of principal components that accounted for at least 80% of the variance among all significantly associated probes. PCA was performed using the scikit-learn package in Python (2.7.17) (47).

### Gene ontology enrichment analysis

We tested whether common CpGs identified across all six marginal linear regression EWAS models were over-represented among gene ontology (GO) terms using the gometh function from the missMethyl R package version 1.34 (48). The probability of significant differential methylation due to the number of probes per gene was taken into consideration. Statistically significant results were defined as having P_FDR_ < 0.05.

### Bayesian EWAS

Probe-by-probe (marginal) linear regression models fail to consider the correlation structure that exists across the methylome. Therefore, we considered Bayesian penalised regression, conducted using BayesR+ (49), as a secondary analysis. This method estimates single marker or probe effects whilst controlling for all other probes as well as being able to control for known and unknown confounding variables. This method also estimates the amount of phenotypic variation attributed to genome-wide DNA methylation. We applied the same covariate and phenotype preparation strategy as in the linear regression models. Significant CpGs were defined as sites with a posterior inclusion probability (PIP) ≥ 0.95. Details on the methods used for the Bayesian strategy can be found in the **Supplementary Methods**.

### Replication of previous literature

The EWAS catalogue (16) was used to determine if the overlapping CpGs that were found to be associated with all six metabolic traits in the linear regression EWASs have previously been identified in other studies. The EWAS catalogue was filtered to whole blood samples, CpG-metabolic trait associations with P < 3.6 × 10^-8^ (in line with our study and consistent with Saffari, *et al* (46)) and study sample *N* > 1,000 participants. The search terms used to identify traits from the EWAS catalogue can be found in **Supplementary Table 2**. The EWAS catalogue was not filtered for studies that may contain GS data.

### Generation and projection of DNA methylation-based proxies of six metabolic traits

Penalised regression models were trained in GS to generate epigenetic scores (EpiScores) of each of the six metabolic traits using the R package biglasso (version 1.5.2). Each trait was modelled as the response variable (using the same phenotype files from the EWASs) and 395,380 CpGs (the 450K methylation array subset that was present in GS) were used as predictors. Cross-validation was carried out (n_folds_ = 20) and an elastic net (elnet) penalty was set (alpha = 0.5). CpG sites with a non-zero coefficient were retained and used to derive EpiScores in LBC1936 (n = 861). This was followed by further testing in the HELIOS cohort (n = 2,245). Missing CpGs were mean imputed in LBC1936 and HELIOS. Predictors obtained from the Bayesian penalised regression models were also projected into LBC1936 and HELIOS using the mean posterior effect sizes as weights for the scores. The variance explained (incremental R^2^) in each metabolic trait by their corresponding EpiScore over and above age and sex in linear regression models was then calculated. In HELIOS, the variance explained was calculated in the full cohort and ancestry subgroups. In HELIOS full cohort models, additional adjustments for ancestry were included.

### EpiScore associations with general cognitive function and change in LBC1936

A latent intercept and age-related slope for general cognitive function were generated in LBC1936 using a structural equation modelling (SEM) framework with the R package Lavaan (version 0.6.12) (50). Measured traits and EpiScores were regressed on intercepts and slopes in separate linear models. Full details are provided in **Supplementary Methods** and **Supplementary Tables 3-6**.

## Results

### Epigenome Wide Association Studies (EWASs) of six metabolic traits

Correlations between metabolic traits in GS ranged between -0.36 (WHR and HDL cholesterol) and 0.6 (BMI and body fat percentage), and are shown in **Supplementary Figure 1**. Marginal linear regression EWASs of six metabolic traits were performed in GS. The number of CpG sites significantly associated (P < 3.6 × 10^-8^) with each of the traits are summarised in **Table 2**. This ranged between 304 for glucose to 11,815 for BMI. Manhattan plots can be observed in **Supplementary Figure 2** and the top 1,000 significantly associated CpGs with each trait are listed in **Supplementary Table 7**. Full summary statistic output will be available upon publication.

**Table 2:**
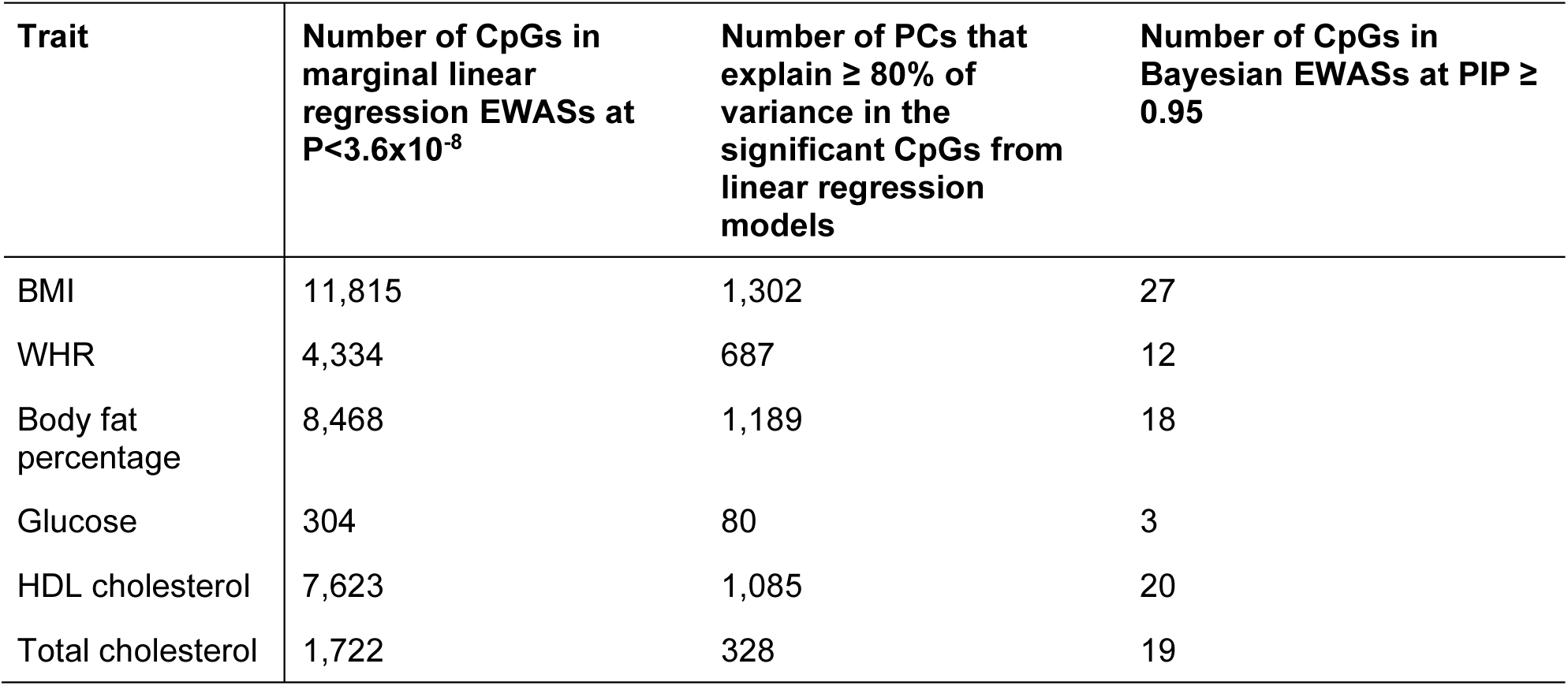
The number of significantly associated CpGs with each metabolic trait in Generation Scotland. The table shows the number of significantly associated CpGs with each metabolic trait using marginal linear regression and Bayesian penalised regression. The table also shows the number of principal components that account for ≥ 80% of the variance of the significant CpGs from the linear regression analyses for each metabolic trait. BMI = body mass index, WHR = waist-hip ratio, HDL = high-density lipoprotein, PCs = principal components.

The large number of significant associations observed in our models may reflect correlation structures among CpG sites (Quantile-Quantile plots and inflation factors – which ranged between 1.18 and 2.48 – can be observed in **Supplementary Figure 3** and **Supplementary Table 8**). Therefore, we performed PCA for each trait to determine the approximate number of independent features present among CpG sites that surpassed the epigenome-wide significance threshold (P<3.6 × 10^-8^). We identified between 80 and 1,302 (for glucose and BMI, respectively) principal components or ‘independent features’ that accounted for ≥ 80% of the variance in the underlying CpG sites (**Table 2**).

Next, we performed Bayesian penalised regression, which jointly models all CpGs and accounts for genome-wide correlation patterns. **Table 2** shows the number of high-confidence associations (PIP ≥ 0.95), which ranged between 3 (glucose) and 27 associations (BMI) (**Supplementary Table 9**). Using the Bayesian method, we obtained estimates for the variance captured by genome-wide DNA methylation that ranged between 24% for WHR and 53% for BMI (**Supplementary Table 10**).

37 CpG sites were significant (P < 3.6×10^-8^) across all six metabolic traits in the marginal linear regression models (**Supplementary Table 11, Supplementary Figure 4**. In the Bayesian models, a single CpG site, “cg06500161” (mapped to the *ABCG1* gene), was associated with BMI, body fat percentage, HDL cholesterol, total cholesterol, and WHR (PIP ≥ 0.95, **Supplementary Table 9**).

14 of the 37 common CpGs from the linear models had been previously associated with metabolic traits in studies using whole blood samples at P < 3.6 × 10^-8^ and study N > 1000 reported in the EWAS catalogue (**Supplementary Table 11**). Of the 37 CpGs associated with all traits in the linear models, four mapped to the *CPT1A* gene, four mapped to the *ABCG1* gene, and three mapped to the PHGDH gene. Seven of the overlapping CpGs did not map to any genes. The remaining 19 CpGs mapped to unique genes giving a total of 22 unique genes containing the overlapping CpGs. Gene ontology (GO) enrichment analysis of the 37 common CpGs was performed. Eleven GO terms were found to be enriched, including cholesterol biosynthetic process and regulation of lipid storage. The full list of enriched GO terms identified can be found in **Supplementary Table 12**.

### Epigenetic Scores (EpiScores) of metabolic traits tested in the LBC1936 and HELIOS

EpiScores for each of the six metabolic traits were trained in GS using elastic net (elnet) penalised regression and projected into the LBC1936 and HELIOS cohorts. We explored how much additional variance could be accounted for in each metabolic trait by the corresponding EpiScore over and above linear regression models adjusting for age and sex. In the LBC1936, EpiScores accounted for 3.2% of the variance for total cholesterol, 18.5% for HDL cholesterol, and 14.4% of the variance in BMI. In HELIOS full cohort analysis, the incremental R^2^ estimates ranged between 7.1% (for total cholesterol) to 20.8% (for BMI). However, there was variability within the ancestry-specific subsets of HELIOS. Most notably, the body fat percentage EpiScore accounted for 9.2% and 9.5% in the Chinese and Malay subgroups but only 3.1% in the Indian subgroup (**Figure 2, Supplementary Table 13**). In LBC1936 and HELIOS, the correlations between all six EpiScores are shown in **Supplementary Figure 5**. Correlations between measured traits ranged from -0.3 – 0.38 for LBC1936, and -0.46 - 0.47 for HELIOS (**Supplementary Figure 6**). Correlations between measured traits and EpiScores ranged between -0.41 – 0.5 in LBC1936 and -0.66 – 0.92 in HELIOS (**Supplementary Figure 7)**.

**Figure 2:**
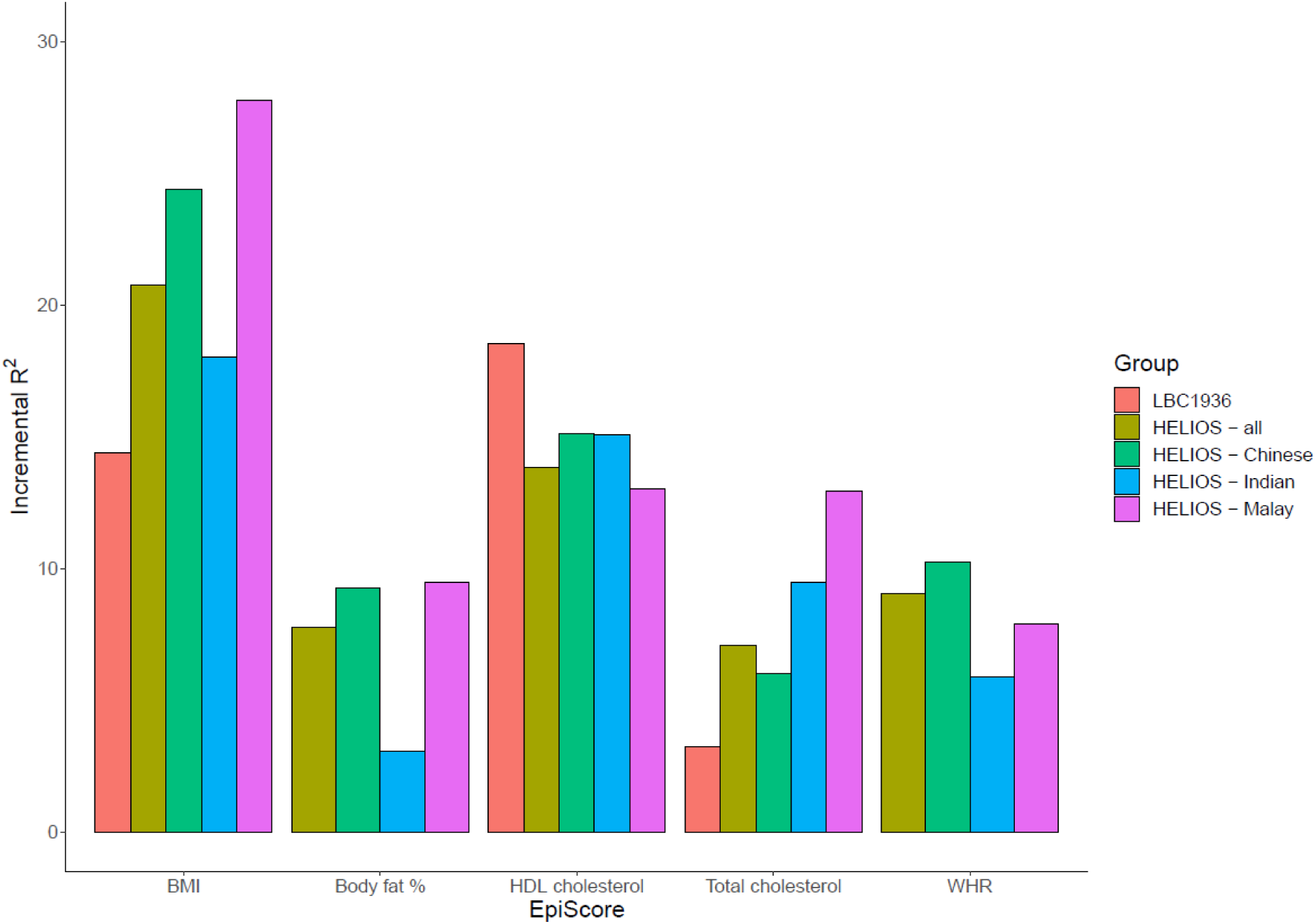
The variance explained in measured metabolic traits by elnet EpiScores in the Lothian Birth Cohort 1936 (LBC1936) and the Health for Life in Singapore (HELIOS) study. Additional variance (incremental R^2^) accounted for in each metabolic trait by their corresponding elnet EpiScores over and above age and sex-adjusted (and ancestry in HELIOS full cohort) linear regression models in LBC1936 and HELIOS. Measured glucose levels were not available for either cohort. Incremental R^2^ was calculated for each ancestry group and in the whole cohort in HELIOS. BMI = body mass index, WHR = waist-hip ratio, HDL = high-density lipoprotein.

Next, we tested the Bayesian EpiScores in both LBC1936 and HELIOS, observing similar results to the elnet approach (**Supplementary Figure 8, Supplementary Table 13**).

### EpiScore associations with general cognitive function

Metabolic traits have previously been linked to cognitive outcomes. Given this, we tested if the metabolic (elnet) EpiScores were associated with general cognitive function level and longitudinal changes in the LBC1936 (n=861). In models adjusting for age and sex, the three measured traits (BMI, total cholesterol and HDL cholesterol) and all EpiScores, except the total cholesterol EpiScore, were significantly associated with general cognitive function (intercept) in LBC1936 (P_FDR_ < 0.05, **Supplementary Figure 9, Supplementary Table 14)**. In fully-adjusted models, significant (P_FDR_ < 0.05) EpiScore associations were observed for WHR, glucose, body fat percentage and BMI (standardized β_range_ -0.08 to -0.12), and for measured BMI (standardized β: - 0.10, **Figure 3A**). No significant associations were observed with general cognitive change over ∼12 years (mean age 70 to mean age 82) of follow-up (P_FDR_ > 0.05, **Supplementary Table 14**). A combination of EpiScore and measured trait accounted for more variance in general cognitive function level than EpiScore or measured trait alone (**Figure 3B, Supplementary Table 15**). EpiScores augmented the measured trait variance explained for general cognitive function by an average of 0.3%.

**Figure 3:**
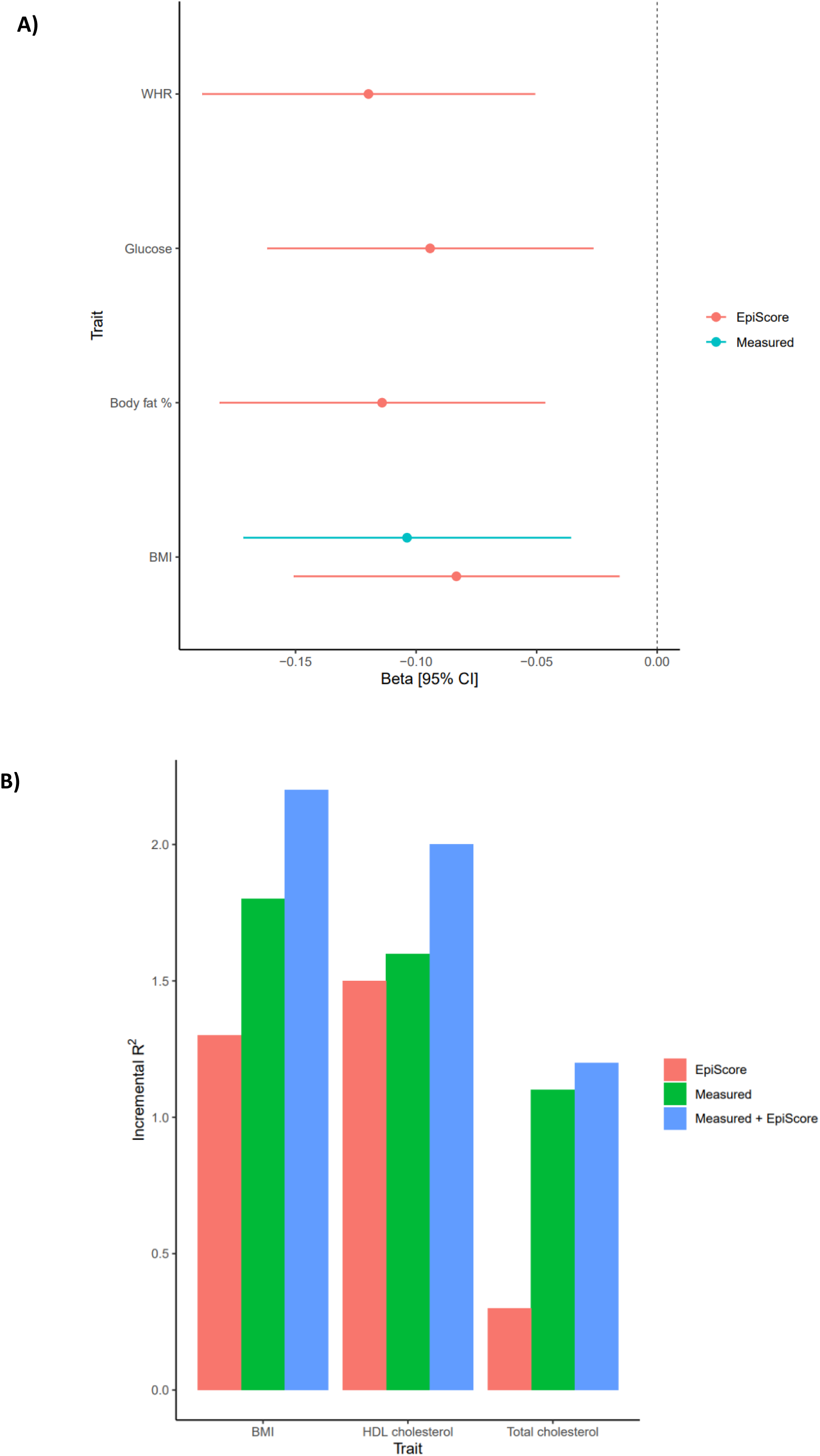
EpiScore and measured metabolic trait in relation to general cognitive function level in the Lothian Birth Cohort 1936 (LBC1936). Panel A shows the significant (PFDR < 0.05) associations between measured traits/EpiScores and general cognitive function level in models with full adjustments. Error bars represent 95% confidence intervals. Panel B shows the additional variance accounted for in general cognitive function level by measured metabolic traits, metabolic Episcores and both combined, over and above linear regression models adjusted for age and sex.

## Discussion

Epigenome-wide association studies of six metabolic traits were performed in Generation Scotland (N > 17,303). A large number of significantly associated CpGs were identified for each trait via linear regression (marginal associations with P < 3.6 × 10^-8^ ranged from 304 to 11,815 per trait). A Bayesian approach, which modelled the CpGs jointly and conditionally upon each other, resulted in between 3 and 27 high confidence (PIP ≥ 0.95) CpG associations for the six traits. EpiScores for each metabolic trait were trained in GS and projected into two independent test cohorts, LBC1936 and HELIOS. The metabolic EpiScores were tested for associations with general cognitive function level and change. Four of the EpiScores were associated with general cognitive function in fully adjusted models (P_FDR_ < 0.05), but none were associated with longitudinal cognitive change.

37 CpGs were associated with all six traits when using the marginal linear regression modelling approach. This included 14 CpGs previously linked to metabolic traits in the literature (17–24, 32, 51–54). Gene ontology analysis revealed the genes that the overlapping CpGs mapped to were enriched for relevant biological functions, including regulation of lipid storage, and cholesterol biosynthetic process. Several genes the 37 CpGs mapped to had known metabolic functions. *ABCG1* and *ABCA1* are part of the ABC transporter superfamily involved in the transport of cholesterol (55, 56). *CPT1A* is a rate-limiting fatty acid oxidation enzyme that oxidises medium and long acyl-CoA esters, an important step that allows these molecules access to the inner mitochondrial membrane (57). *PDK4* is a kinase that inhibits the pyruvate dehydrogenase complex (PDC) which is responsible for the decarboxylation of pyruvate to acetyl-CoA (58). The inhibition of *PDC* results in a switch from glucose oxidation to fatty-acid oxidation and *PDK4* has been suggested as a marker for increased fatty-acid oxidation (58, 59).

Metabolic EpiScores accounted for additional variance in metabolic traits over and above age and sex in both LBC1936 and HELIOS. The elnet EpiScores for BMI and total cholesterol accounted for more variance in their corresponding measured traits in the HELIOS full cohort than in the LBC1936. Conversely, the EpiScore for HDL cholesterol accounted for more variance in the LBC1936 than in the HELIOS full cohort. The performance of elnet metabolic EpiScores in HELIOS varied by ancestry group. In particular, the body fat percentage EpiScore performed similarly in Chinese and Malay individuals (∼9% variance accounted for) but had a much lower performance in Indian participants (3.1% variance accounted for). Within the Asian population, it has been reported that Indians have a higher body fat percentage compared with Chinese and Malay populations (60). Asian Indian individuals also have been shown to have increased total and centrally distributed body fat compared with those of European ancestries (61).

The potential usefulness of using DNA methylation to impute measured traits in studies where they are not available was highlighted by the similarity of effect sizes between metabolic EpiScores and their corresponding measured traits in models predicting general cognitive function level (basic adjustments).

This study has multiple strengths including large sample sizes, the use of multi-ancestry cohorts, a multi-method approach (linear regression and Bayesian penalised regression), volunteers from a wide range of ages across adulthood, and longitudinal data to test for cognitive changes in late-life testing (LBC1936). Of the two EWAS strategies, and despite adjustments for relevant covariates, the marginal linear regression approach yielded a vast number of significant CpGs associated with each metabolic trait. However, this approach is naïve in that it does not account for the genome-wide correlation patterns and structure across the methylome. This leads to an inflation in the number of significant findings and biased estimation of effect sizes.Using more stringent methods like BayesR+ helped to overcome such issues, resulting in a high confidence set of CpG-trait associations. Another key strength of the study is that the metabolic EpiScores trained in a cohort of individuals residing in Scotland could account for variance in metabolic traits in a multi-ancestry cohort of Chinese, Malay and Indian Singaporeans. A limitation is that only three of the six metabolic traits were measured in LBC1936, therefore we were unable to compare EpiScore performance against measured WHR, glucose and body fat percentage in this cohort. Finally, alternative strategies for feature pre-selection prior to training EpiScores are likely to result in improved predictors (62, 63).

To conclude, our findings suggest that different EWAS strategies (i.e., marginal linear models and conditional Bayesian models) vastly alter the number of significant CpGs associated with metabolic traits. As increasingly large cohorts with DNA methylation are generated, conditional analyses will help to control false positive rates although they will not identify all correlated/co-dependent sites under a peak. We have also shown that metabolic EpiScores trained in a Scottish population perform well in external Scottish and multi-ancestry Singaporean cohorts. However, further testing is required in e.g., populations from African or Hispanic ancestries to determine how well the predictors generalise. Further, metabolic EpiScores and measured metabolic traits had comparable magnitudes of association with general cognitive function. This highlights the potential usefulness of metabolic EpiScores to “impute” the corresponding traits where they have not been measured in a cohort.

## Availability of data and material

According to the terms of consent for Generation Scotland participants, access to data must be reviewed by the Generation Scotland Access Committee. Applications should be made to.

Lothian Birth Cohort data are available on request from the Lothian Birth Cohort Study, University of Edinburgh (https://www.ed.ac.uk/lothian-birth-cohorts/data-access-collaboration). Lothian Birth Cohort data are not publicly available due to them containing information that could compromise participant consent and confidentiality.

HELIOS data are available on request from the study’s principal investigators. Data access requests for this study should be directed to

All code associated with this manuscript is available for open access at the following GitHub repository: https://github.com/hmsmith22/metabolic_trait_project

EWAS summary statistics will be submitted to the EWAS Catalog and Edinburgh DataShare upon publication.

## Ethics approval and consent to participate

All components of GS received ethical approval from the NHS Tayside Committee on Medical Research Ethics (REC Reference Number: 05/S1401/89). GS has also been granted Research Tissue Bank status by the East of Scotland Research Ethics Service (REC Reference Number: 20-ES-0021), providing generic ethical approval for a wide range of uses within medical research.

Ethical approval for the LBC1936 study was obtained from the Multi-Centre Research Ethics Committee for Scotland (Wave 1, MREC/01/0/56) and the Lothian Research Ethics committee (Wave 1, LREC/2003/2/29) and the Scotland A Research Ethics Committee (Waves 2-5, 07/MRE00/58). All participants provided written informed consent. These studies were performed in accordance with the Helsinki declaration.

The HELIOS study was approved by the National Technological University (NTU) Institutional Review Board [IRB-2016-11-030], with written informed consent obtained from each participant before the commencement of the study.

## Consent for publication

Not applicable

