## Supplementary Figures for "Methylome-wide studies of six metabolic traits"

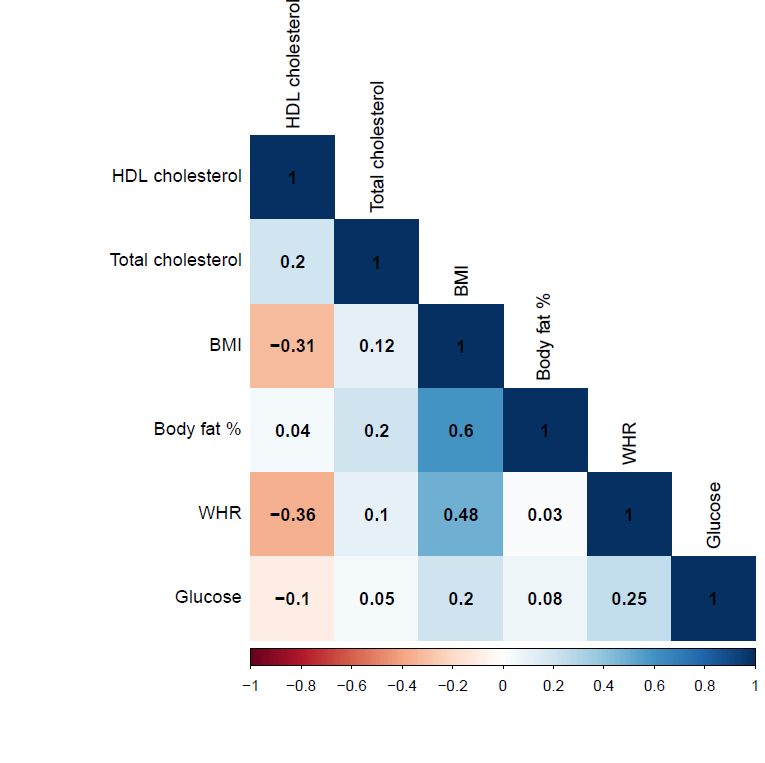

**Supplementary Figure 1: Metabolic trait correlations in Generation Scotland.** The heatmap shows the phenotypic Pearson correlations between the metabolic traits in Generation Scotland. BMI = body mass index; WHR = waist-hip ratio; HDL = high-density lipoprotein.

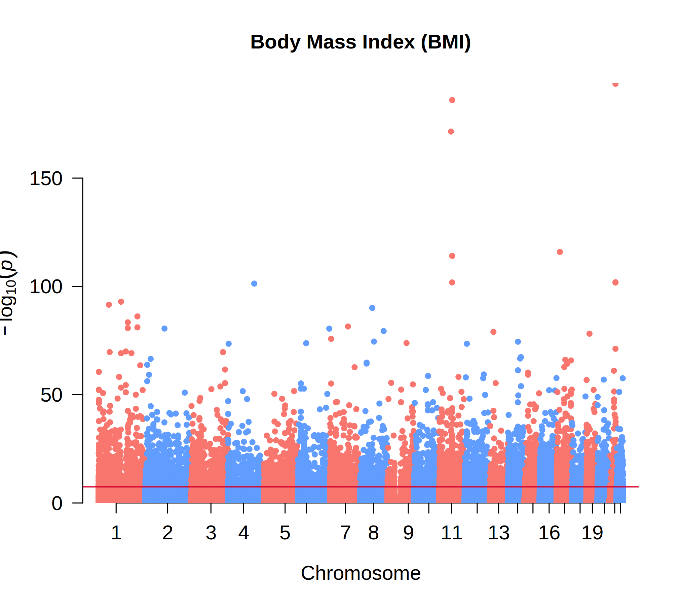

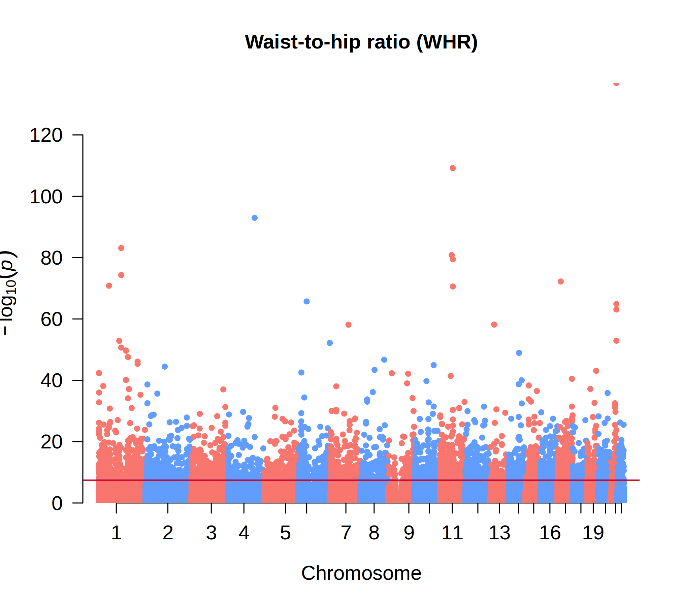

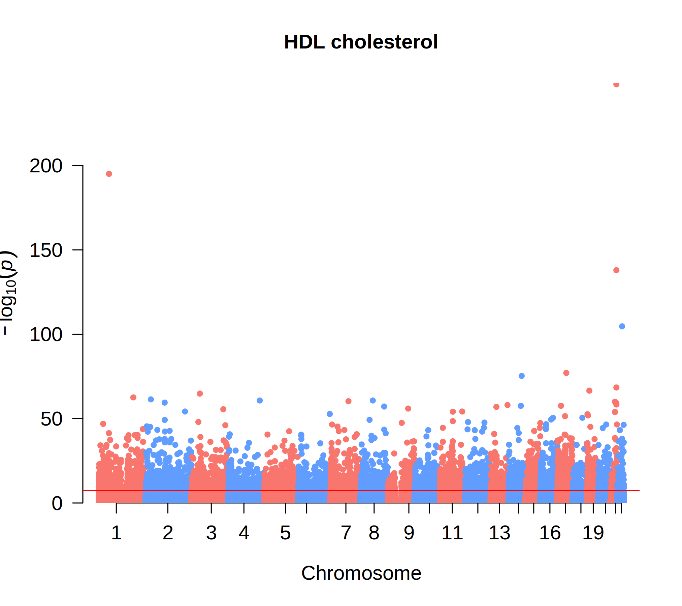

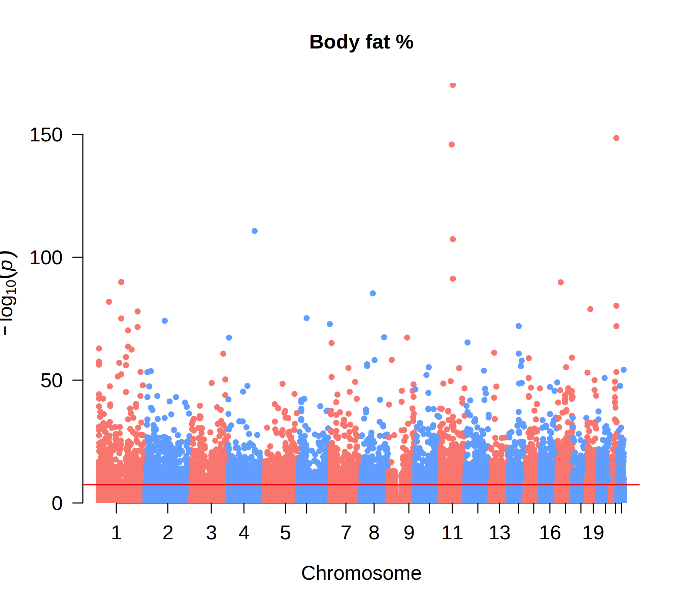

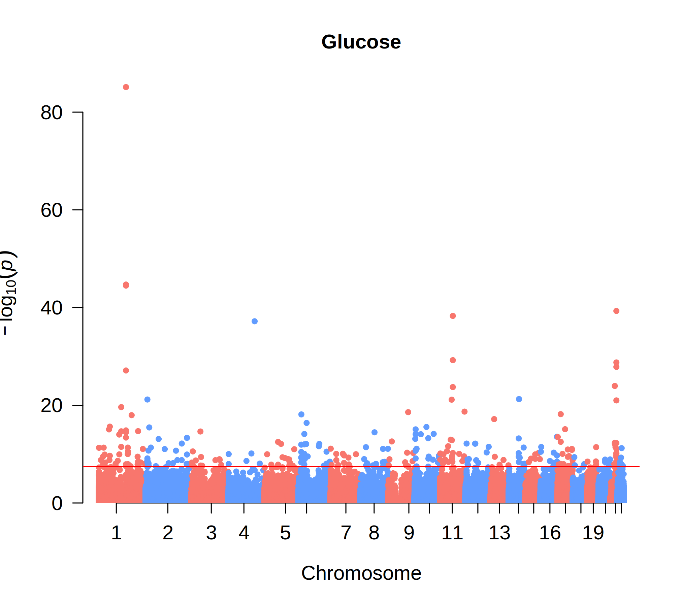

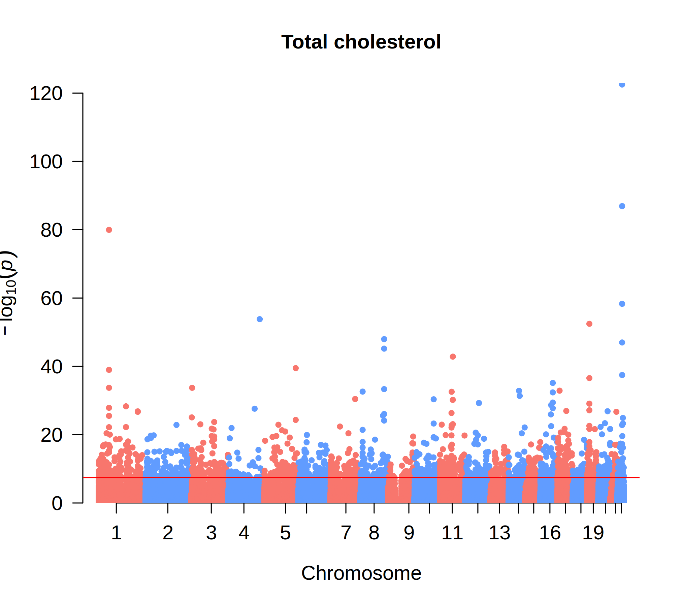

**Supplementary Figure 2: Manhattan plots of the results from the marginal linear regression epigenome-wide association studies of six metabolic traits in Generation Scotland.** The Manhattan plots for each of the six metabolic traits show each CpG as a data point. The x-axis shows the chromosome position, and the y-axis shows the association significance (-log_10_(P)) for each CpG site. The horizontal red line indicates the significance threshold (P < 3.6 x 10^-8^). BMI = body mass index; WHR = waist-hip ratio; HDL = high-density lipoprotein.

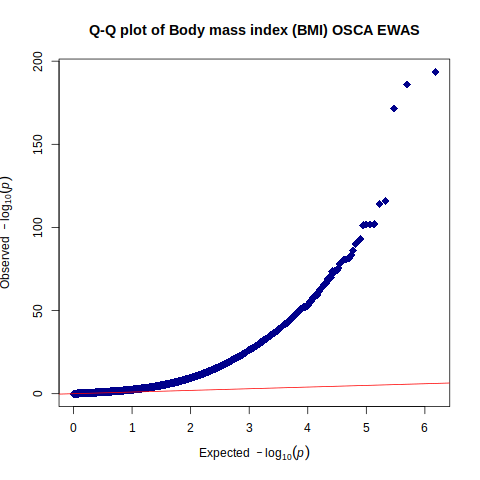

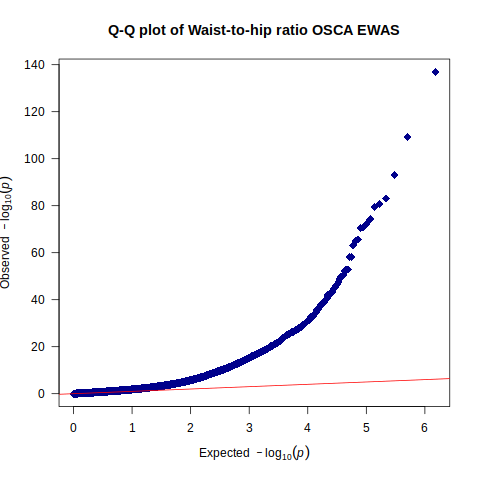

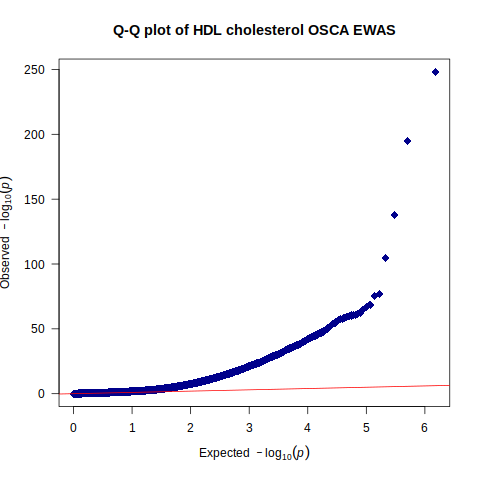

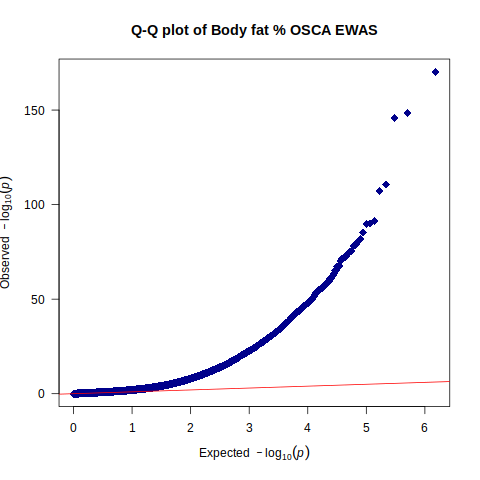

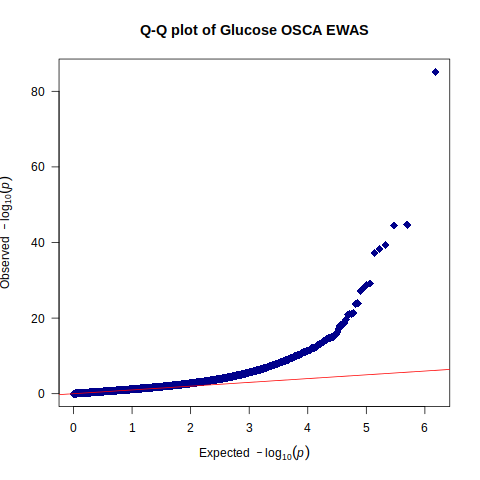

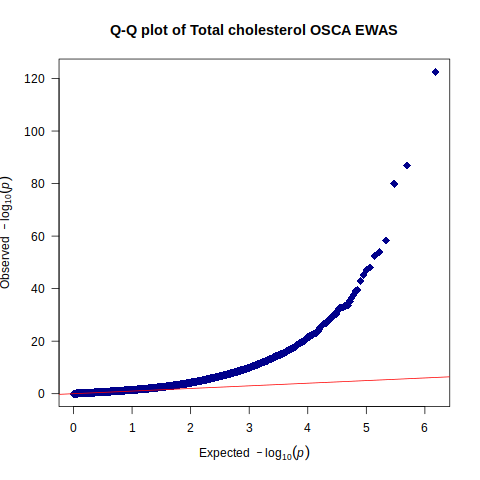

**Supplementary Figure 3: Quantile-Quantile plots of the results from the marginal linear regression epigenome-wide association studies of six metabolic traits in Generation Scotland.** The plots show expected –log_10_(P) by the observed –log_10_(P) for each metabolic trait. The red line shows a trend line of where the observed and expected values are the same. BMI = body mass index; WHR = waist-hip ratio; HDL = high-density lipoprotein.

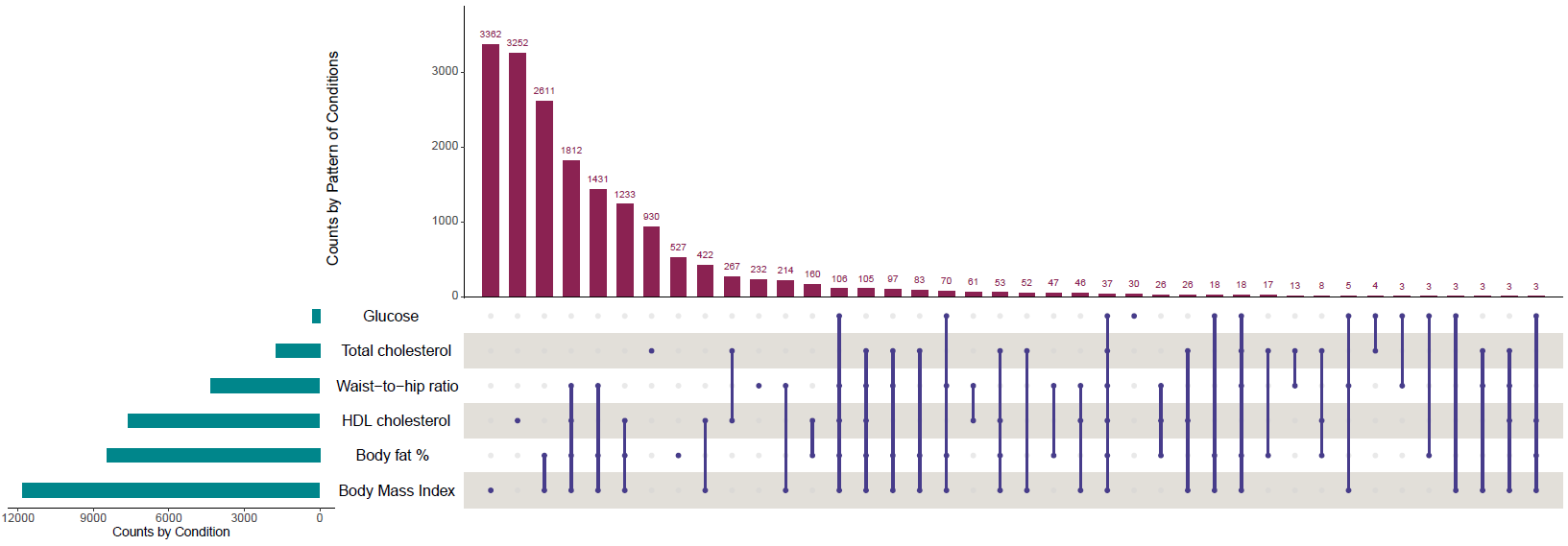

**Supplementary Figure 4: An upset plot of significant CpGs for the six metabolic traits in Generation Scotland.**  The figure shows the number of unique and overlapping CpGs for the six metabolic traits from the marginal linear regression models. BMI = body mass index; WHR = waist-hip ratio; HDL = high-density lipoprotein.

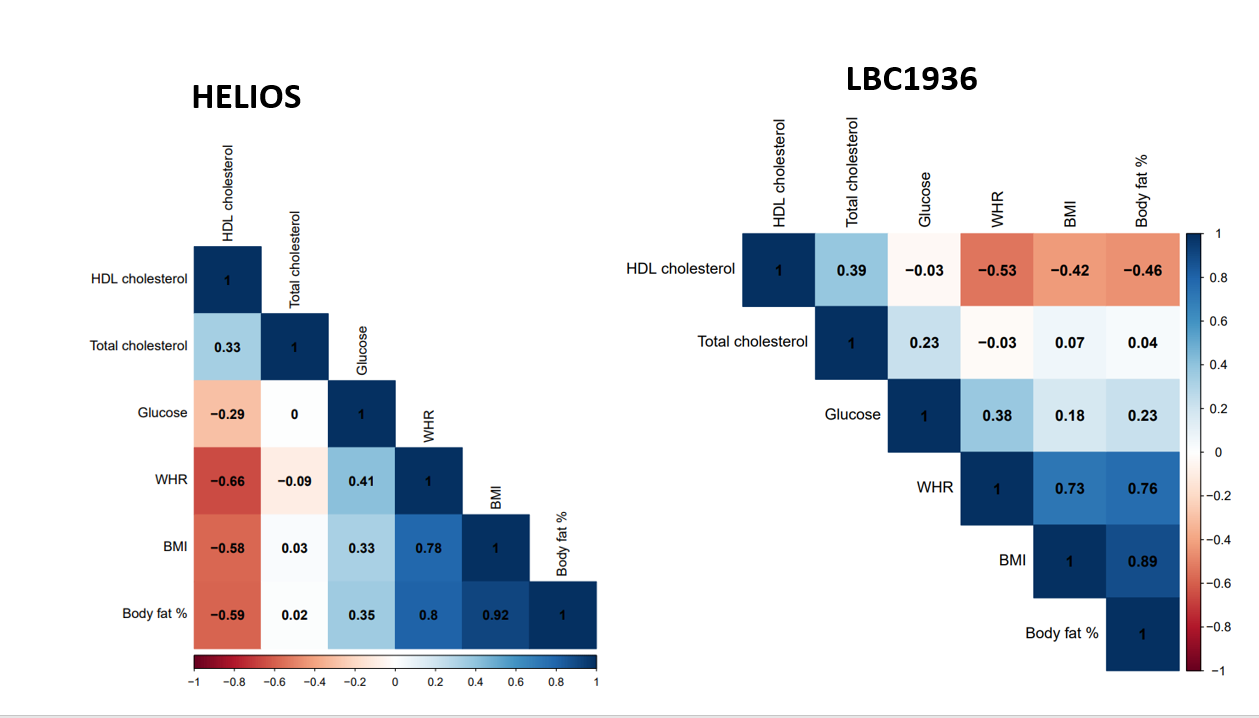

**Supplementary Figure 5: Metabolic EpiScore correlations for the Health for Life in Singapore (HELIOS) study and the Lothian Birth Cohort 1936 (LBC1936).** The heatmaps show the Pearson correlation between each metabolic EpiScore in HELIOS and LBC1936. BMI = body mass index; WHR = waist-hip ratio; HDL = high-density lipoprotein.

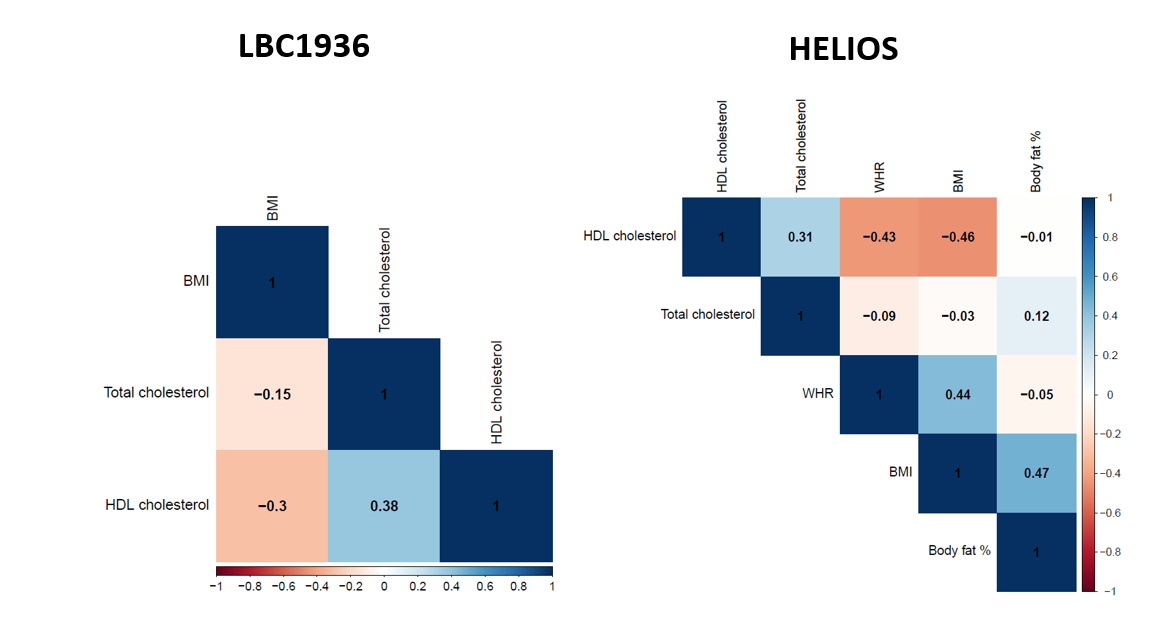

**Supplementary Figure 6: Metabolic trait correlations in the Health for Life in Singapore (HELIOS) study and the Lothian Birth Cohort 1936 (LBC1936).** The heatmaps show the Pearson correlation between measured metabolic traits in HELIOS and LBC1936. BMI = body mass index; WHR = waist-hip ratio; HDL = high-density lipoprotein.

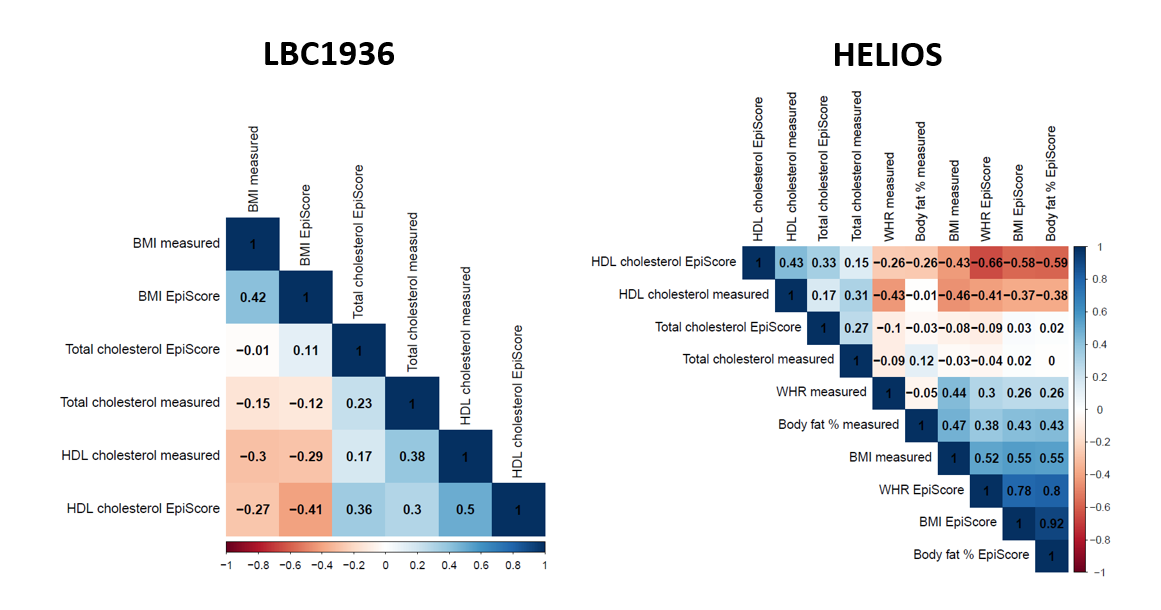

**Supplementary Figure 7: EpiScore-metabolic trait correlations in the Health for Life in Singapore (HELIOS) and the Lothian Birth Cohort (LBC1936).** The heatmaps show the Pearson correlations between metabolic EpiScores and measured metabolic traits in HELIOS and LBC1936. BMI = body mass index; WHR = waist-hip ratio; HDL = high-density lipoprotein.

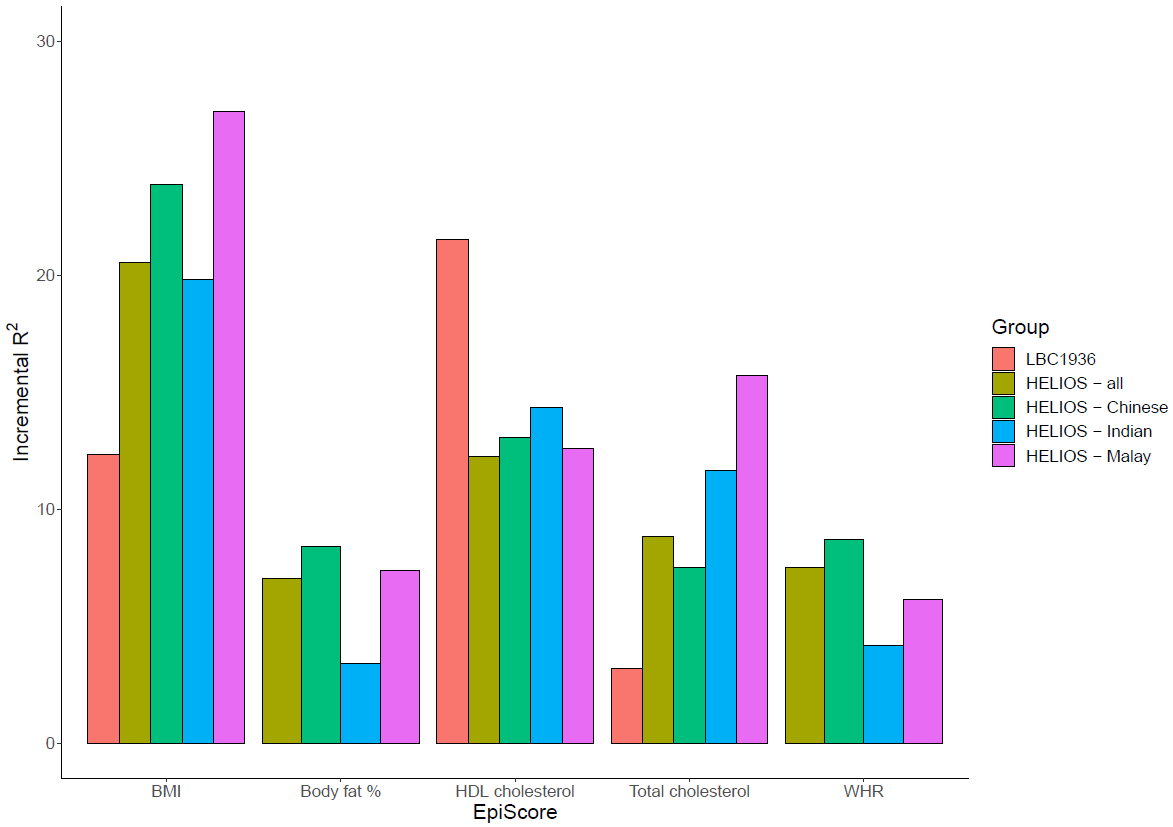
**Supplementary Figure 8: The variance explained in measured metabolic traits by Bayesian EpiScores in the Health for Life in Singapore (HELIOS) study and the Lothian Birth Cohort 1936 (LBC1936).** The figure shows the incremental R^2^ for each metabolic trait accounted for by their corresponding Bayesian metabolic EpiScores over and above age and sex-adjusted linear regression models in LBC1936 and HELIOS. The incremental R^2^ was calculated for each ancestry group and in the whole cohort for the HELIOS study. Full cohort models in HELIOS were additionally adjusted for ancestry. BMI = body mass index; WHR = waist-hip ratio; HDL = high-density lipoprotein.

.

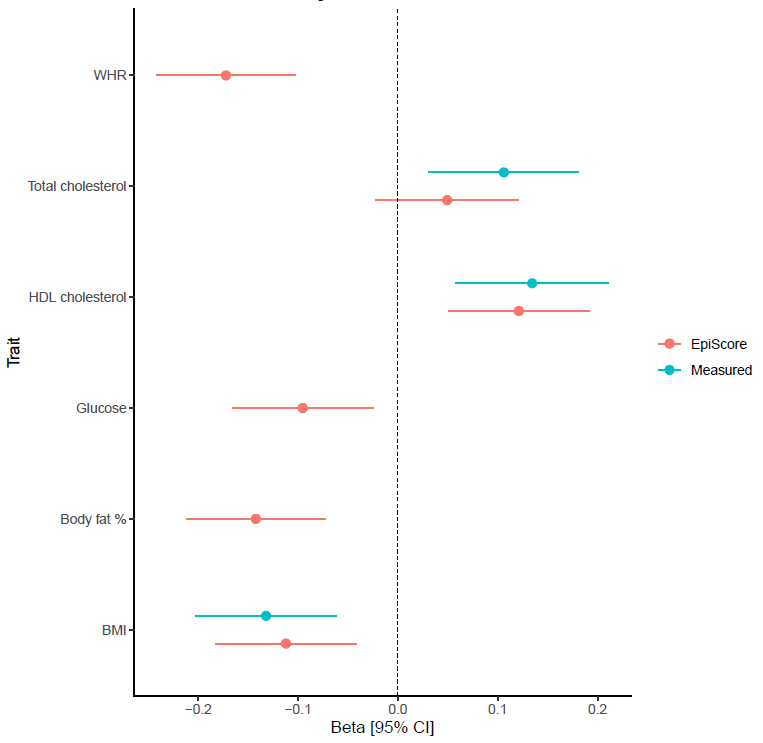

**Supplementary Figure 9:** **EpiScore and measured metabolic trait associations with general cognitive function level in the Lothian Birth Cohort (LBC1936).** The figure shows associations between measured metabolic traits/EpiScores and general cognitive function level in models with basic adjustments (age and sex). The error bars represent 95% confidence intervals. BMI = body mass index; WHR = waist-hip ratio; HDL = high-density lipoprotein.
