## Supplementary Methods for "Methylome-wide studies of six metabolic traits"

***Metabolic measures in Generation Scotland (GS), Lothian Birth Cohort 1936 (LBC1936) and the Health for Life in Singapore (HELIOS) study***

This study investigated six metabolic measures including body mass index (BMI), waist-hip ratio (WHR), body fat percentage, high-density lipoprotein (HDL) cholesterol, total cholesterol and glucose. The Generation Scotland (GS) study had all six traits available for analysis. The HELIOS study had all traits available for analysis except glucose. The LBC1936 had only BMI, HDL cholesterol and total cholesterol available for analysis. Outlier removal strategies were chosen on a cohort-by-cohort basis in line with previous approaches. In GS, BMI, body fat percentage and WHR outliers were removed by visual inspection after bivariate plots of all pairwise combinations (n_removed_ = 426). Outliers > 4 standard deviations from the mean were removed for glucose, HDL cholesterol and total cholesterol (n_removed_ = 173, 26, 15, respectively). In LBC1936, measured metabolic trait data were visually inspected and no outliers were removed. In HELIOS, data points were considered outliers if they were beyond 3.5 standard deviations from the mean. BMI (weight in kg /height in m^2^), WHR (waist/hip circumference) and body fat percentage were measured in the clinic. In HELIOS, whole-body DEXA scans were used to quantify body fat (1). In GS, body fat percentage is quantified with bioimpedance. HDL cholesterol, total cholesterol and glucose from blood samples were measured in mmol/L.

***DNA methylation in Generation Scotland***

The GS cohort consists of 3 sets of participants: N_set 1_= 5,087, N_set 2_ = 4,450, N_set 3_ = 8,876, and 121 experimental batches. The Illumina Methylation EPIC (850K) array was used to quantify DNA methylation. A subset of set 1 were related individuals determined by family ID. Participants in set 2 were not related to each other or related to the individuals in set 1. Set 3 included some participants that were related to each other or related to participants in set 2/3.

Details of quality control (QC) performed have been published previously (2-4). Briefly, outliers were removed based on the log median intensity of the methylated vs unmethylated signal per array. Probes were also removed if in more than 5% of samples, the bead count was < 3 or the p-value > 0.05. If probes belonged to the sex chromosomes, or if they overlay any SNPs and/or resided in a probable cross-hybridising location they were removed. If the predicted sex and recorded sex did not match samples were removed. Samples were removed if they didn’t match the expected sample type (i.e., were not blood), if ≥ 1% of CpGs had a detection p-value > 0.05, and/or if a participant responded “yes” to all self-reported diseases in questionnaires. Dasen normalisation was carried out across all individuals. Once QC was complete, 752,722 CpGs for 18,411 individuals were available for analysis.

***DNA methylation in the Lothian Birth Cohort 1936***

In LBC1936, blood-based DNA methylation was measured using the Illumina methylation array (450K). DNAm QC has been described previously (5-7). Briefly, the data was background-corrected and normalised to controls. Samples were removed for several reasons including probes with low detection and call rates (<95% and detected < 450,000 at P < 0.01), incorrect DNAm predicted sex and genotype-SNP mismatches. 459,310 CpGs for 861 individuals were available for analysis after QC.

***DNA methylation in Health for Life in Singapore (HELIOS)***

DNA methylation in the HELIOS cohort was measured using the Illumina HumanMethylation EPIC array after Bisulfite conversion of DNA was carried out according to the manufacturer’s protocol (EZ DNA methylation kit). The *minfi* software package (8) was used to obtain bead intensity with the detection rate of P < 0.02 used for marker calling. Probes with call rates <95% were excluded. Samples were excluded for array scanning failures (n = 2), incorrect DNAm predicted sex (n = 39), and duplication (n = 17). 2,445 samples with 837,722 CpG sites were available for analysis after QC. Quantile normalisation was used to account for batch effects.

***Bayesian EWAS***

BayesR+ is a software implemented in C++ for performing Bayesian penalised regression on complex continuous traits (9). A prior distribution is assumed as a mixture of Gaussian distributions, which correspond to groups of probes with different effect sizes. A discrete spike at zero is included, which removes probes that have a negligible effect on the trait. Informed by data from a previous analysis of BMI, prior mixture variances of probes were set to 0.0001, 0.001, 0.01 (9). Pre-corrected phenotype and DNA methylation data were scaled to mean zero and unit variance. Gibbs sampling was used to sample over the posterior distribution and consisted of 10,000 samples with 5,000 as burn-in. A thinning of 5 samples was applied to reduce autocorrelation. Four chains were used and the final 250 samples per chain (after thinning) were combined to form the final set of 1,000 iterations from which variance and effect size estimates were obtained. Probes with a posterior inclusion probability (PIP) ≥ 95% were deemed to be significant.

**Cognitive measures in the Lothian Birth Cohort 1936 (LBC1936)**

Cognitive measures in the LBC1936 have been described previously (10-13). Cognitive testing was repeated for 5 waves at ages 70, 73, 76, 79, and 82. Thirteen cognitive measures for all five waves were available. Several cognitive domains were measured including visuospatial ability (tests: Block Design, Matrix Reasoning (WAIS-III^UK^) and Spatial span (WMS-III^UK^)), memory (tests: Verbal Paired Associates, Logical Memory – a combination of immediate and delayed memory (WMS-III^UK^) and Digit-span backwards (WAIS-III^UK^)), and verbal ability (tests: National Adult Reading Test, Wechsler Adult Reading Test and Verbal Fluency Test (using letters V, F and L)). Processing speed was evaluated using the Digit-symbol, Symbol Search (WAIS-III^UK^), Choice Reaction Time and Inspection Data availability and descriptive statistics for each measure can be found in **Supplementary Table 3**.

**General cognitive function level and change in LBC1936**

Latent measures of general cognitive function and change were generated using confirmatory factor analysis in a structural equation modelling (SEM) framework using the *Lavaan* (version 0.6-12) R package (14). Intercepts and slopes of each cognitive test were used to indicate a latent intercept and slope (level and change) of general cognitive function (**Supplementary Table 4**). Levels and changes in cognitive functioning were modelled with a latent growth curve model (LGCM) using a Factor of Curves specification (15). A first-order hierarchical structure was specified, and residual covariance between tests in the same cognitive domain was included, in line with a previously established correlational structure of cognitive domains (speed, memory, verbal ability and visuospatial (16)). Residual covariance between intercept and slope for individual tests was also modelled. Within-wave residual covariance between the National Adult Reading Test and the Wechsler Adult Reading Test were modelled as these tests were highly correlated. The marker method was used to scale according to the first variable to aid model convergence. Negative latent residual variances were fixed to zero. Full information maximum likelihood was used to include all data available. Confirmatory factor index (CFI), Tucker-Lewis index (TLI), root mean squared error approximation (RMSEA) and the standardised root mean squared residual (SRMR) fit measures are reported (**Supplementary Table 5**). Linear regression models were run in Lavaan to test associations between general cognitive function level and change, and the metabolic traits/EpiScores with basic- and full-adjustments as follows:

*Basic model: Latent G factor (intercept or slope) ~ measured trait/EpiScore + Age at baseline + Sex*

*Full model: Latent G factor (intercept or slope) ~ measured trait/EpiScore + Age at baseline + Sex + Scottish Index of Multiple Deprivation (SIMD) + Epigenetic smoking score (EpiSmokEr) + Alcohol units per week*

Descriptive statistics for each covariate in LBC1936 can be found in **Supplementary Table 6**.
